# Genetic Liability to Higher Muscle Strength Associates with a Lower Risk of Cardiovascular Disease Mortality in Men Irrespective of Physical Activity in Adulthood: A Longitudinal Cohort Study

**DOI:** 10.1101/2024.05.31.24308268

**Authors:** Päivi Herranen, Katja Waller, Laura Joensuu, Teemu Palviainen, Eija K Laakkonen, Jaakko Kaprio, Elina Sillanpää

## Abstract

**Background:** Low muscle strength predicts premature mortality. We determined whether genetic liability to muscle strength is associated with mortality and whether this association is influenced by long-term leisure-time physical activity (PA).

**Methods and Results:** We estimated the effects of a polygenic score for hand grip strength (PGS HGS) on all-cause and cardiovascular disease (CVD) mortality risk in the older Finnish Twin Cohort (N=8815, 53% women). National registries provided dates and causes of death. PA volume was assessed longitudinally in 1975, 1981, and 1990 using validated questionnaires. During the 16.9-year median follow-up time (143,723 person-years), 2896 deaths occurred, of which 1089 were due to CVD. We found a significant interaction between sex and PGS HGS (*P*=0.016) for predicting all-cause mortality. In men, one standard deviation increase in the PGS HGS was associated with a decreased risk both of all-cause (hazard ratio, HR [95% confidence interval, CI]): 0.93 [0.89–0.98] and CVD mortality (HR 0.88 [0.81–0.96]). Associations persisted after adjusting for PA, but only with CVD mortality after adjusting for other lifestyle covariates (HR 0.85 [0.76–0.96]). The cumulative incidence rates by age 75 years were 4.3% lower for all-cause mortality and 2.1% lower for CVD mortality in the highest PGS HGS quintile compared to the lowest quintile. No PGS HGS×PA interactions were found. PGS HGS was not associated with mortality in women.

**Conclusions:** Higher PGS HGS was associated with a decreased risk of all-cause and CVD mortality in men; however, long-term PA in adulthood did not potentiate this association.

**Clinical Perspective:** *What Is New?:*

- To the best of our knowledge, this is the first study to use a genome-wide polygenic score for hand grip strength to investigate whether the association between genetic liability to muscle strength and lifespan is affected by physical activity.
- Our results suggest that individuals with a genetic predisposition for higher muscle strength have a modest decreased risk of cardiovascular disease mortality, independent of their lifestyle.

*What Are the Clinical Implications?:*

- Polygenic scores for muscle strength require further development but may help identify individuals who represent extreme ends of genetic predisposition and vulnerability to premature death.

## Introduction

Maximal voluntary muscle strength reflects an individual’s intrinsic capacity for physical function.^1^ It starts to diminish gradually from around the third decade of life, with a 1–2% decline annually after reaching 50 years of age.^2^ The loss of muscle strength and mass is influenced by a combination of variables, such as genetic factors, lifestyle choices, disease burden, random effects, and natural aging processes.^3^ Preserving adequate strength is crucial to maintaining the mobility and health of older individuals. Notably, higher muscle strength, especially handgrip strength (HGS), is categorically established as a predictor of lower risk of cardiovascular outcomes^4^, including all-cause and cardiovascular disease (CVD) mortality.^5,6^

The association between low muscle strength and premature mortality can presumably be explained by the increased disability and risk for traumatic events such as injuries and fractures,^7^ resulting from a loss of muscle function. However, emerging evidence suggests the additional pathways, through increased risk of CVD and related risk factors,^5,6^ pulmonary diseases, and cancers.^6^ While other measures of muscle quantity or quality, such as thigh circumference appear to be associated with increased risk of all-cause and CVD mortality,^8^ recent evidence indicates that muscle strength is a stronger predictor of future adversities than, for example, muscle mass.^3^ Individual patterns of muscle strength remain highly stable as evidenced by strong correlations between measurements taken from middle age to old age decades apart,^9^ with high accuracy in predicting future health outcomes even in younger populations.^10–12^ This implies that an individual’s trajectory for maximal strength during aging may follow a predictable pattern and describe their intrinsic capacity for lifetime health.^13^

This also highlights the exceptional role of genetics as a significant determinant of maximal muscle strength and associated outcomes over the life course. According to twin studies, heritability estimates for muscle strength vary between 30%–65%.^14^ Previous genome-wide association studies (GWASs) have identified multiple genetic variants, each of which individually has only a small effect on muscle strength, indicating its polygenic nature.^15,16^ Polygenic scores (PGSs) summarize the combined effects of genetic markers on an individual’s genetic predisposition to a particular trait or disease and may be useful in investigating gene-environment (G × E) interplay.^17^ G × E interaction occurs when the expression of genetic predisposition is influenced or altered by environmental effects, or when environmental conditions augment or moderate the effects of a specific genetic variation on a trait.^18^ We recently derived a PGS for HGS (PGS HGS) and established it is a meaningful predictor of variability in overall muscle strength; the PGS HGS explained for 6.1% of the variation in the HGS and 5.4% of the variation in knee extension strength measured among the older Finnish population.^19^ It was also associated with several disability phenotypes, common diseases, and lifespan.^19,20^ However, it is unclear whether these associations are independent of lifestyle, such as physical activity (PA).

PA encompasses any bodily movement involving skeletal muscles, ranging from daily tasks to structured and targeted exercises.^21^ The relationship between muscle strength and PA is multifaceted and mutually reinforcing. There is a bidirectional causal association between the two, which implies that the age-related loss of muscle strength results in reduced PA levels, which accelerates the decline in mobility and increases the risk of functional limitations and disability.^22^ Consequently, the loss of maximal muscle strength due to biological aging can be mitigated by PA.^22,23^ Previous studies have reported that six months of structured interventions for middle-aged and older adults can help regain the muscle strength as much as is typically lost in 10 years of aging.^24,25^ Furthermore, it has been proposed that greater muscle strength may boost a person’s willingness to be physically active.^26^ Numerous studies have also established a robust link between regular PA and reduced mortality risk^21,27,28^ which can be attributed to the beneficial effects of PA, including improvements in cardiorespiratory fitness, increased insulin sensitivity, and glycemic control which are closely interconnected with muscle strength.^21,22,27^ These findings underscore the importance of maintaining muscle strength by engaging in active lifestyles.

In the current study, we used the PGS HGS to examine whether genetic predisposition for higher muscle strength predicts all-cause and CVD mortality. We hypothesized that individuals with a genetic susceptibility to higher muscle strength tend to have a decreased mortality risk, and that higher PA levels could amplify the genetic influence on higher muscle strength, in turn decreasing the risk of premature mortality. The second aim was to determine whether long-term leisure-time PA mediates the relationship between PGS HGS and mortality assuming that individuals with a higher genetic predisposition for higher muscle strength might be more inclined to PA.

## Methods

### Study Design and Participants

The older Finnish Twin Cohort (FTC)^29^ consisted of twin pairs born before 1958, with both twins in the pair alive in 1974. A baseline survey was conducted in 1975, with two follow-up questionnaire surveys administered via mail in 1981 and 1990. The cohort included mortality follow-up data from Finnish national registries. From the FTC data, we extracted data for all genotyped participants aged 18–50 years at the time of the baseline survey of 1975 (N = 8815). Individual PGS HGS were calculated for these participants, and mortality analyses were conducted.

### Research Ethics and Data Access

All data collection procedures were carried out in accordance with the Declaration of Helsinki. The FTC study protocol was approved by the ethics committees of the University of Helsinki and Helsinki University Central Hospital (approval numbers: 113/E3/2001 and 346/E0/05, respectively). All participants were informed about the study, and their written consent was obtained. The full cohort data cannot be publicly available owing to the potential identifiability of the twin siblings in Finland. DNA samples and follow-up questionnaire data are available for the twins who had consented to transfer their data to the Biobank of the National Institute for Health and Welfare; these data are publicly available by following a standardized application procedure (https://thl.fi/en/research-and-development/thl-biobank/for-researchers/application-process). Full cohort data can be obtained from the Institute for Molecular Medicine Finland Data Access Committee (FIMM-DAC) by authorized researchers who have the appropriate IRB/ethics approval and an institutionally approved study plan. The GWAS’s summary statistics for maximum HGS were based on genetic and phenotypic data from the UK Biobank (UKBB) and derived from the publicly accessible website of the Pan-UKBB (https://pan.ukbb.broadinstitute.org/).

### Genotyping and Polygenic Scoring

Genotyping, quality control, and imputation for the UKBB study and the older FTC are described in the UKBB’s documents and previously published studies.^30–33^ The PGS HGS pipeline and validation against measured muscle strength phenotypes in an independent cohort sample have been described by Herranen et al.^19^ Briefly, we used publicly available Pan-UKBB GWAS summary statistics regarding the maximum HGS for 418 827 individuals (restricted to individuals with European ancestry). In the UKBB cohort, the maximum HGS was measured using a calibrated hydraulic hand dynamometer (Jamar J00105, Lafayette Instrument Company, IN, USA). We used a method based on weighing the GWAS summary statistics using a linkage disequilibrium (LD) reference panel and multiple Bayesian regression models (SBayesR).^34^ This methodology possesses better prediction accuracy than other commonly used summary statistic-based methods. The reference sample included 50 000 UKBB participants, who were randomly selected from the original cohort. For computational purposes and to capture variation across the whole genome we restricted genome data to ∼1.1 million HapMap3 single nucleotide polymorphisms (SNPs), which are likely well imputed and common in European samples (MAF > 5%).^35^ The major histocompatibility complex region from chromosome 6 (6p22.1-21.3) was excluded. We used a total of 1 006 473 variants to calculate the PGS in our analysis. In the older FTC, the PGS HGS was computed for each individual by calculating the weighted sum of the risk allele dosages for each variant.

### All-Cause and CVD Mortality

Data regarding the date and cause of death dates were obtained from the population register of Finland and Statistics Finland; deaths were covered until December 31, 2020. The Finnish registries include personal identification numbers permitting record linkage, ensuring high accuracy in determining cause-of-death, death certification, and coding practices.^36,37^ Deaths due to CVD (International Classification of Diseases, ICD-9 codes: 390–459, ICD-10 codes: I00–I99) or coronary heart diseases (ICD-9 codes: 410–414, ICD-10 codes: I20–I25) were classified as CVD mortality. Mortality rates in the older FTC correspond to the general Finnish adult population.^29^

### Lifestyle Covariates

*Long-term leisure-time PA* encompassed leisure-time PA, as well as commuting activities. It was assessed in metabolic equivalent (MET) hours per day (h/day) using a structured questionnaire regarding activity intensity, duration, and frequency, as measured in 1975, 1981, and 1990.^29^ The mean value from available time points was used in the analysis.

Data for self-reported *education years* (in 1981) were categorized as follows: less than primary school (3 years of education), primary school (6 years), junior high school (9 years), high school graduate (12 years), university degrees (16 years), and ≥ 1 year of education such as vocational training in addition to primary school (7 years), junior high school (10 years) or high school (13 years).^38^

*Body mass index* (BMI, kg/m^2^) was calculated based on self-reported height and weight. Self-reported BMI was validated in a previous study.^39^ The latest known BMI from either 1981 or 1990 was used in the analysis.

*Smoking* status was noted on the latest known self-reported information from either 1990 or 1981 and was classified as never, occasional, former, or current smokers. Current smokers were further classified into light (1–9), medium (10–19), and heavy (≥20) smokers as per the number of cigarettes smoked per day.^40^

*Alcohol* use was calculated based on the latest known average alcohol consumption (g/day) from either the 1990 or 1981 survey.^41^

### Statistical Analyses

We assessed associations between standardized PGS HGS and all-cause and CVD mortality via Cox proportional hazards (PH) models using age as the time scale. Each survival model considered left truncation by initiating the follow-up interval from the individual time of DNA sampling (1993–2017); follow-up continued until death, emigration, or end-of-follow-up (December 31, 2020). We verified PH assumptions by inspecting Schoenfeld residuals; in the case of deviation from the PH assumptions, the Cox models were additionally fitted with a time-varying covariate^42^ in age intervals (≤ 65 years, 65–75 years, 75–85 years, and > 85 years) as a sensitivity analysis (Tables S1–S5). Mortality risk was calculated per one standard deviation (SD) change in the PGS HGS. We reported the results as hazard ratios (HRs) with 95% confidence intervals (CIs). To account for genetic stratification occurring in the Finnish population, we adjusted all models for the first 10 genetic principal components (PCs) of ancestry and utilized clustering for the family identifier to address family relatedness in the data. As the interaction term between sex and PGS HGS was statistically significant with all-cause mortality (*P*= 0.016; HR [95% confidence interval, CI]: HR 1.14 [1.03–1.27], and borderline with CVD mortality (*P*= 0.050; HR 1.22 [1.00–1.48], we performed separate analyses for men and women. The first set of models included PGS HGS and 10 genetic PCs, the second set of models was additionally adjusted for PA. The final models incorporated further adjustments for lifestyle and socioeconomic covariates known to be associated with premature mortality, based on previous literature. To investigate PA as a possible moderator in the relationship between PGS HGS and mortality, we replicated the analysis by incorporating the interaction term PGS HGS × PA into the final model. Furthermore, we estimated potential mediation pathways by analyzing the association between PGS HGS and PA using linear mixed models adjusted for age in 1990 and 10 genetic PCs. Finally, the cumulative incidence of all-cause and CVD mortality by age at 75 years was estimated by dividing PGS HGS into quintiles (low: < 20%, intermediate: 20–80%, and high: > 80%). In addition, we assessed the discrimination ability of each predictor and possible improvements in mortality risk prediction with and without PGS HGS among men in the complete cases (no missing data) models by inspecting the concordance index (c-index). As diseases may influence PA levels, we also conducted sensitivity analysis among apparently healthy participants (no physician-diagnosed or self-reported or inpatient admissions, or use of reimbursable medication for selected chronic cardiometabolic diseases and cancers in 1981).^43^

All analyses were conducted using R software (version 4.2.3); statistical significance was set at *P* < 0.05.

## Results

Among 8815 participants a total of 2896 deaths occurred, of which 1089 were CVD-related; 37% of the men and 29% of the women died during the median follow-up time of 16.9 years (143 723 person-years). A summary of the participants’ baseline characteristics is presented in Table 1.

**Table 1.**
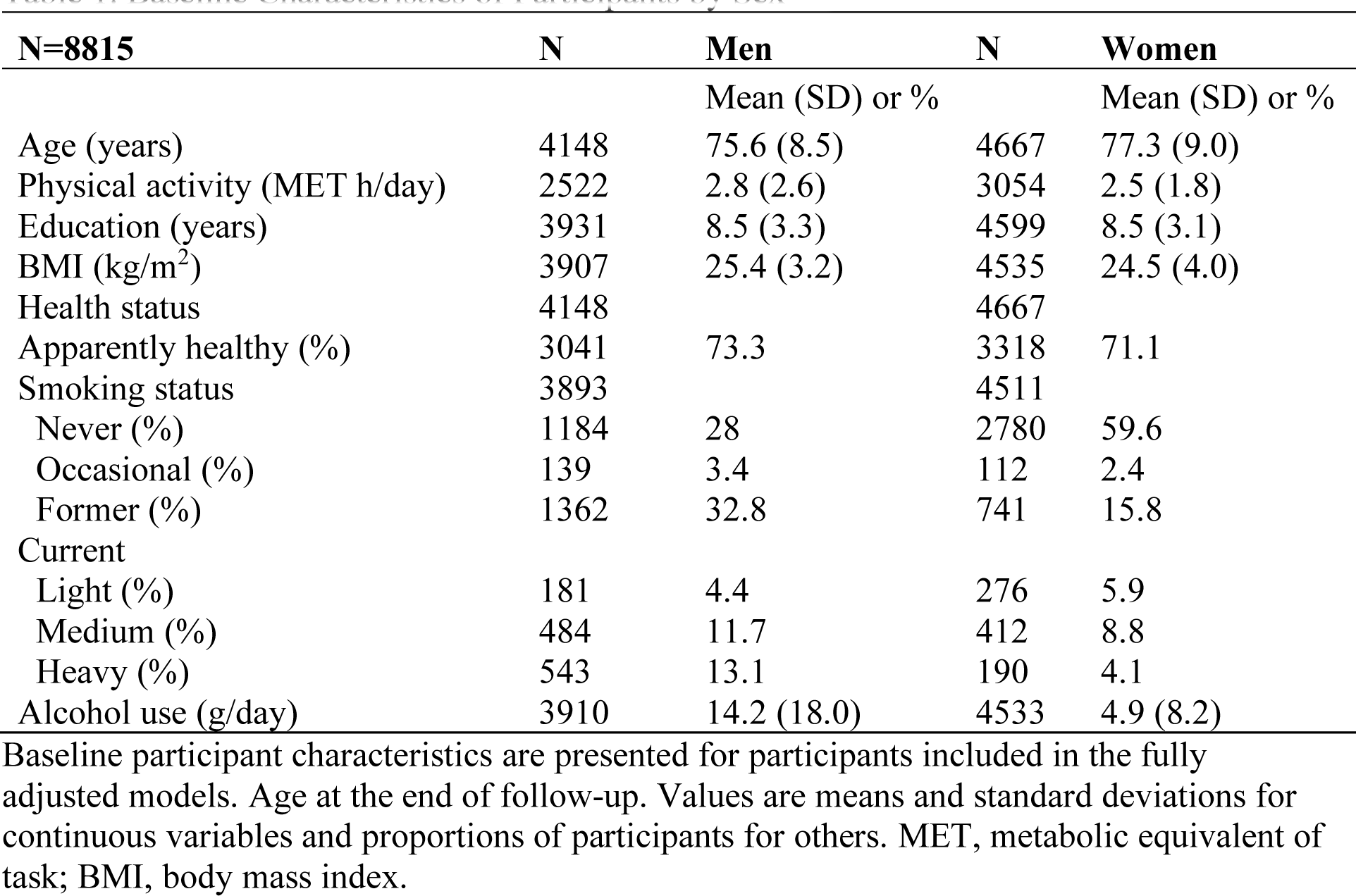
Baseline Characteristics of Participants by Sex.

| **N=8815** | **N** | **Men** | **N** | **Women** |
| --- | --- | --- | --- | --- |
|  |  | Mean (SD) or % |  | Mean (SD) or % |
| Age (years) | 4148 | 75.6 (8.5) | 4667 | 77.3 (9.0) |
| Physical activity (MET h/day) | 2522 | 2.8 (2.6) | 3054 | 2.5 (1.8) |
| Education (years) | 3931 | 8.5 (3.3) | 4599 | 8.5 (3.1) |
| BMI (kg/m<sup>2</sup>) | 3907 | 25.4 (3.2) | 4535 | 24.5 (4.0) |
| Health status | 4148 |  | 4667 |  |
| Apparently healthy (%) | 3041 | 73.3 | 3318 | 71.1 |
| Smoking status | 3893 |  | 4511 |  |
| Never (%) | 1184 | 28 | 2780 | 59.6 |
| Occasional (%) | 139 | 3.4 | 112 | 2.4 |
| Former (%) | 1362 | 32.8 | 741 | 15.8 |
| Current |  |  |  |  |
| Light (%) | 181 | 4.4 | 276 | 5.9 |
| Medium (%) | 484 | 11.7 | 412 | 8.8 |
| Heavy (%) | 543 | 13.1 | 190 | 4.1 |
| Alcohol use (g/day) | 3910 | 14.2 (18.0) | 4533 | 4.9 (8.2) |

Baseline participant characteristics are presented for participants included in the fully adjusted models. Age at the end of follow-up. Values are means and standard deviations for continuous variables and proportions of participants for others. MET, metabolic equivalent of task; BMI, body mass index.

Tables 2–5 summarize the results of the associations between PGS HGS and mortality. In men, higher PGS HGS was associated with lower all-cause and CVD mortality HR [95% confidence interval, CI]: HR 0.93 [0.89–0.98] and HR 0.88 [0.81–0.96], respectively; Tables 2 and 3). Four out of 10 (41 %) men in the lowest PGS HGS quintile died during the follow-up period, compared with 34% in the highest quintile (HR 0.81 [0.69–0.95]). Furthermore, the cumulative incidence rates of all-cause and CVD mortality in the highest PGS HGS quintile were slightly lower than those in the lowest quintile (Figures 1 and 2). For instance, by 75 years of age, the incidence of all-cause mortality in the highest PGS HGS quintile was 4.3% lower than that in the lowest quintile (survival probability HR 0.73 [0.68–0.77] versus HR 0.69 [0.65–0.74]); likewise, CVD mortality in the highest PGS HGS quintile was 2.1% lower than that in the lowest quintile (HR 0.90 [0.87–0.93] versus HR: 0.88 [0.85–0.91]). In women, PGS HGS was not associated with either all-cause mortality (HR 1.01 [0.96–1.07]) or CVD mortality (HR 0.96 [0.87–1.05]; Tables 4 and 5).

**Figure 1.**
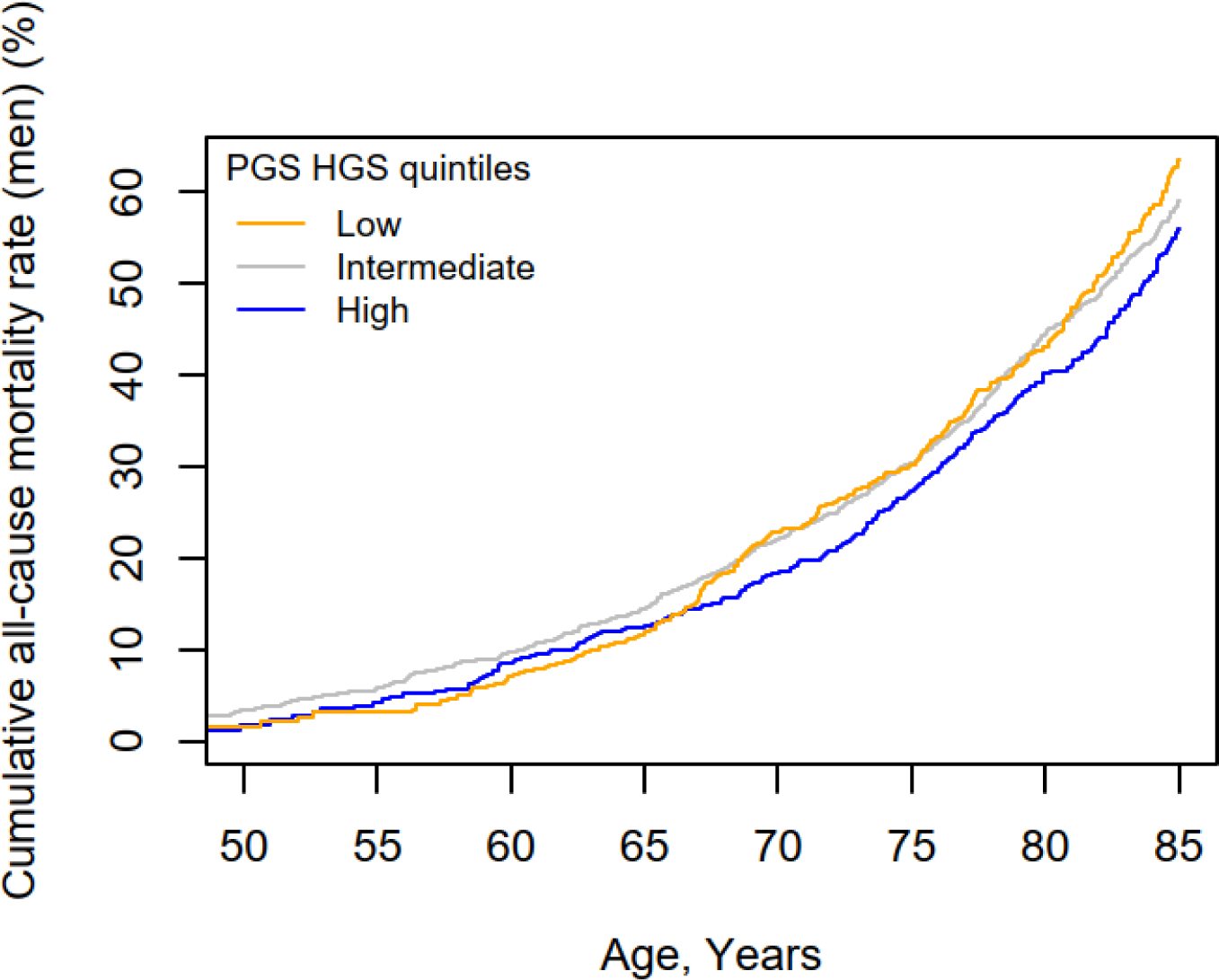
Cumulative incidence of all-cause mortality by PGS HGS categories for the scaled age in men. Cumulative incidence is presented as a percentage. The survival curves are from the Cox regression analysis. Adjusted for 10 genetic principal components of ancestry. PGS HGS, the polygenic score for hand grip strength.

**Figure 2.**
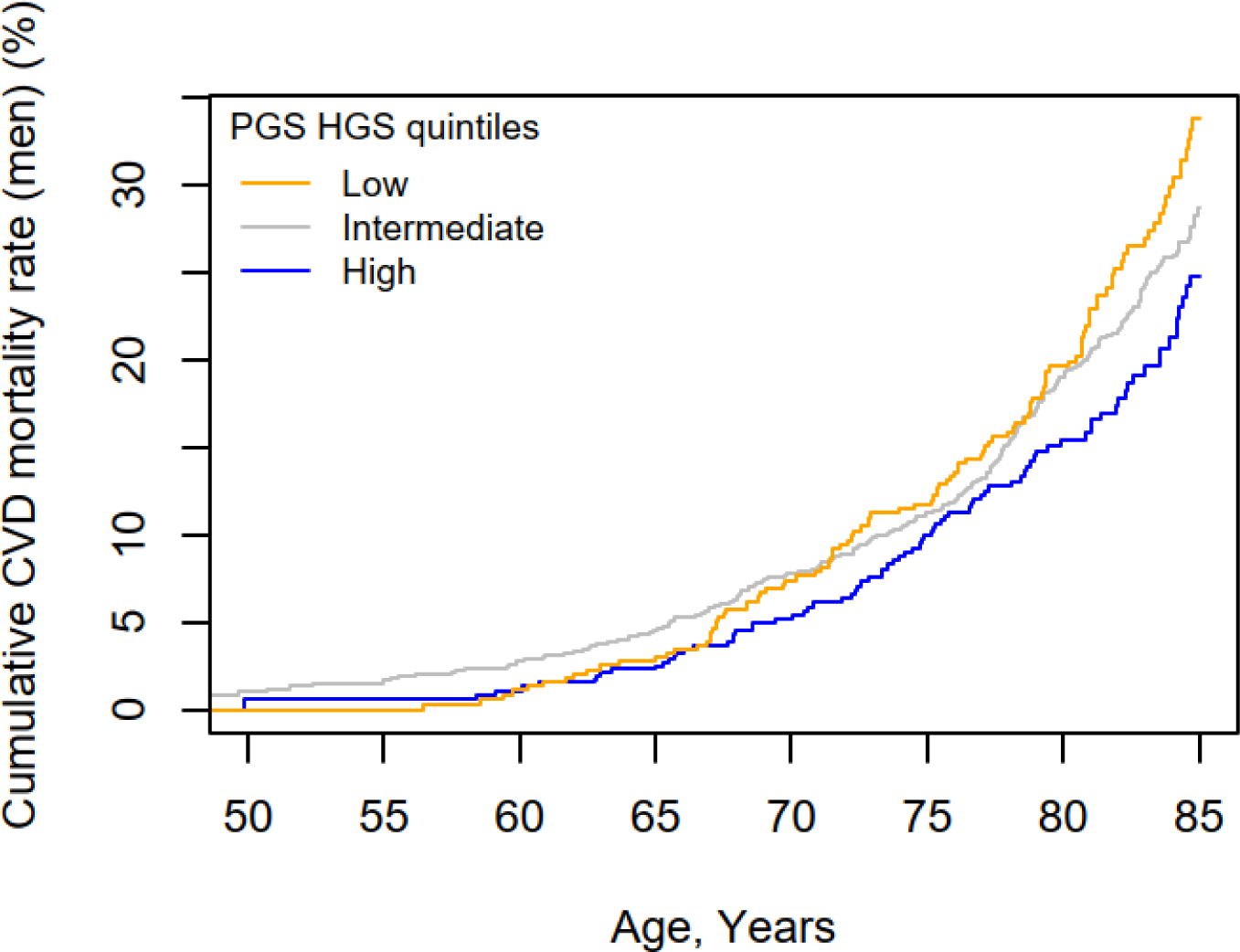
Cumulative incidence of CVD-related mortality by PGS HGS categories for the scaled age in men. Cumulative incidence is presented as a percentage. The survival curves are from the Cox regression analysis. Adjusted for 10 genetic principal components of ancestry. CVD, cardiovascular disease; PGS HGS, the polygenic score for hand grip strength.

**Table 2.**
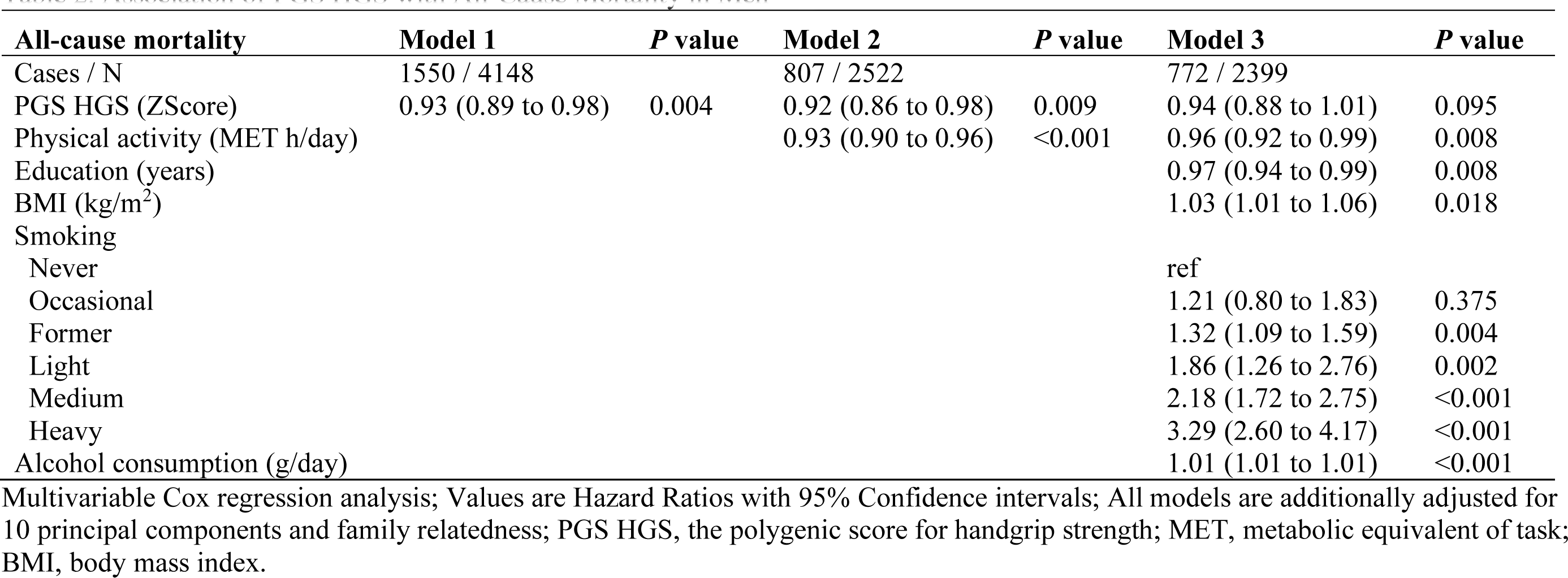
Association of PGS HGS with All-Cause Mortality in Men.

**Table 3.**
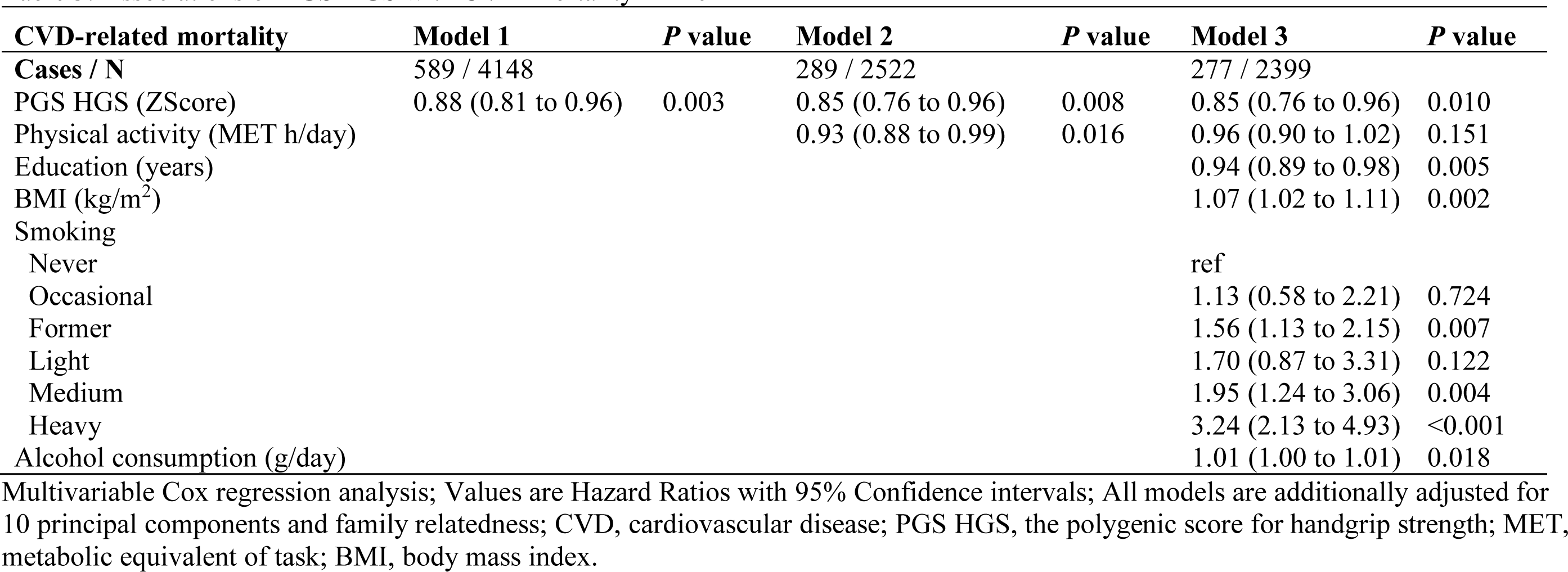
Associations of PGS HGS with CVD Mortality in Men.

| **CVD-related mortality** | **Model 1** | ***P* value** | **Model 2** | ***P* value** | **Model 3** | ***P* value** |
| --- | --- | --- | --- | --- | --- | --- |
| **Cases / N** | 589 / 4148 |  | 289 / 2522 |  | 277 / 2399 |  |
| PGS HGS (ZScore) | 0.88 (0.81 to 0.96) | 0.003 | 0.85 (0.76 to 0.96) | 0.008 | 0.85 (0.76 to 0.96) | 0.010 |
| Physical activity (MET h/day) |  |  | 0.93 (0.88 to 0.99) | 0.016 | 0.96 (0.90 to 1.02) | 0.151 |
| Education (years) |  |  |  |  | 0.94 (0.89 to 0.98) | 0.005 |
| BMI (kg/m<sup>2</sup>) |  |  |  |  | 1.07 (1.02 to 1.11) | 0.002 |
| Smoking |  |  |  |  |  |  |
| Never |  |  |  |  | ref |  |
| Occasional |  |  |  |  | 1.13 (0.58 to 2.21) | 0.724 |
| Former |  |  |  |  | 1.56 (1.13 to 2.15) | 0.007 |
| Light |  |  |  |  | 1.70 (0.87 to 3.31) | 0.122 |
| Medium |  |  |  |  | 1.95 (1.24 to 3.06) | 0.004 |
| Heavy |  |  |  |  | 3.24 (2.13 to 4.93) | <0.001 |
| Alcohol consumption (g/day) |  |  |  |  | 1.01 (1.00 to 1.01) | 0.018 |

Multivariable Cox regression analysis; Values are Hazard Ratios with 95% Confidence intervals; All models are additionally adjusted for 10 principal components and family relatedness; CVD, cardiovascular disease; PGS HGS, the polygenic score for handgrip strength; MET, metabolic equivalent of task; BMI, body mass index.

**Table 4.** Associations of PGS HGS with All-Cause Mortality in Women.

| **All-cause mortality** | **Model 1** | ***P* value** | **Model 2** | ***P* value** | **Model 3** | ***P* value** |
| --- | --- | --- | --- | --- | --- | --- |
| Cases / N | 1346 / 4667 |  | 598 / 3054 |  | 593 / 3012 |  |
| PGS HGS (ZScore) | 1.01 (0.96 to 1.07) | 0.621 | 1.08 (0.99 to 1.17) | 0.082 | 1.08 (0.99 to 1.17) | 0.089 |
| Physical activity (MET h/day) |  |  | 0.99 (0.94 to 1.04) | 0.076 | 1.00 (0.96 to 1.05) | 0.875 |
| Education (years) |  |  |  |  | 0.98 (0.95 to 1.01) | 0.172 |
| BMI (kg/m<sup>2</sup>) |  |  |  |  | 1.04 (1.02 to 1.06) | <0.001 |
| Smoking |  |  |  |  |  |  |
| Never |  |  |  |  | ref |  |
| Occasional |  |  |  |  | 1.30 (0.78 to 2.15) | 0.318 |
| Former |  |  |  |  | 1.23 (0.95 to 1.59) | 0.123 |
| Light |  |  |  |  | 1.41 (0.95 to 2.10) | 0.088 |
| Medium |  |  |  |  | 2.03 (1.53 to 2.70) | <0.001 |
| Heavy |  |  |  |  | 3.70 (2.60 to 5.27) | <0.001 |
| Alcohol consumption g/day |  |  |  |  | 1.01 (1.00 to 1.02) | 0.007 |

Multivariable Cox regression analysis; Values are Hazard Ratios with 95% Confidence intervals; All models are additionally adjusted for 10 principal components and family relatedness; PGS HGS, the polygenic score for handgrip strength; MET, metabolic equivalent of task; BMI, body mass index.

**Table 5.**
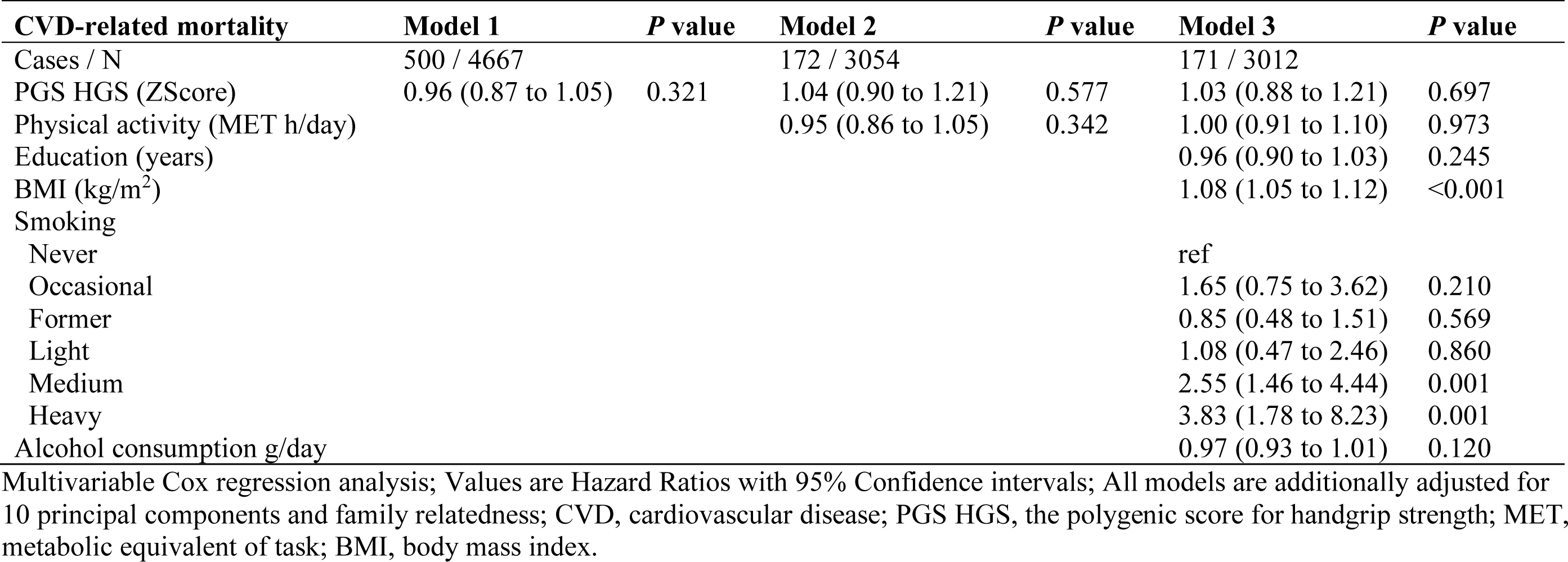
Associations of PGS HGS with CVD Mortality in Women.

| **CVD-related mortality** | **Model 1** | ***P* value** | **Model 2** | ***P* value** | **Model 3** | ***P* value** |
| --- | --- | --- | --- | --- | --- | --- |
| Cases / N | 500 / 4667 |  | 172 / 3054 |  | 171 / 3012 |  |
| PGS HGS (ZScore) | 0.96 (0.87 to 1.05) | 0.321 | 1.04 (0.90 to 1.21) | 0.577 | 1.03 (0.88 to 1.21) | 0.697 |
| Physical activity (MET h/day) |  |  | 0.95 (0.86 to 1.05) | 0.342 | 1.00 (0.91 to 1.10) | 0.973 |
| Education (years) |  |  |  |  | 0.96 (0.90 to 1.03) | 0.245 |
| BMI (kg/m<sup>2</sup>) |  |  |  |  | 1.08 (1.05 to 1.12) | <0.001 |
| Smoking |  |  |  |  |  |  |
| Never |  |  |  |  | ref |  |
| Occasional |  |  |  |  | 1.65 (0.75 to 3.62) | 0.210 |
| Former |  |  |  |  | 0.85 (0.48 to 1.51) | 0.569 |
| Light |  |  |  |  | 1.08 (0.47 to 2.46) | 0.860 |
| Medium |  |  |  |  | 2.55 (1.46 to 4.44) | 0.001 |
| Heavy |  |  |  |  | 3.83 (1.78 to 8.23) | 0.001 |
| Alcohol consumption g/day |  |  |  |  | 0.97 (0.93 to 1.01) | 0.120 |

Multivariable Cox regression analysis; Values are Hazard Ratios with 95% Confidence intervals; All models are additionally adjusted for 10 principal components and family relatedness; CVD, cardiovascular disease; PGS HGS, the polygenic score for handgrip strength; MET, metabolic equivalent of task; BMI, body mass index.

In men, the association of PGS HGS persisted with both all-cause (HR 0.92 [0.86–0.98]) and CVD (HR 0.85 [0.76–0.96]) mortality when models were adjusted for PA (Tables 2 and 3). After further adjustments for lifestyle and socioeconomic covariates, the estimated all-cause mortality risk did not change, although it was no longer statistically significant (HR 0.94, [0.88–1.01]). Results did not change for CVD mortality (HR 0.85 [0.75–0.96]). PA showed independent main effects with all-cause and CVD mortality only in men (HR 0.93 [0.90– 0.96] and HR 0.93 [0.88–0.99], respectively). After further adjustments, this association persisted only for all-cause mortality (HR 0.96 [0.92–0.99]). There was no interaction between PGS HGS and PA (*P*=0.370; Table S6). In all, PGS HGS did not associate with physical activity (in men, β = −0.031, standard error (SE) = 0.052, *P* = 0.551, n =2522, and in women, β = 0.042, SE = 0.037, *P* = 0.195, n =3054, respectively).

The discrimination ability of each predictor was evaluated among men with complete case data (n = 2399 cases; mortality events – all-cause: n = 772; CVD: n = 277). PGS HGS alone exhibited modest predictive ability for both all-cause (c-index [95% CI]): (0.54 [0.53–0.55]) and CVD (0.56 [0.54–0.58]) mortality (Figures 3 and 4). However, the c-indices of PGS HGS were comparable to those of models including other variables – BMI, education, and PA: c-indices ranging between 0.53 and 0.55 for all-cause mortality; and PA, education, and alcohol consumption: c-indices ranging between 0.56 and 0.57 for CVD mortality. In all-cause mortality, the c-index was 0.65 [0.64–0.67] for the model including only lifestyle factors, and 0.66 [0.64–0.67] for the complete model. For CVD mortality, the c-indices were 0.66 [0.63–0.68] and 0.66 [0.64–0.69], respectively.

**Figure 3.**
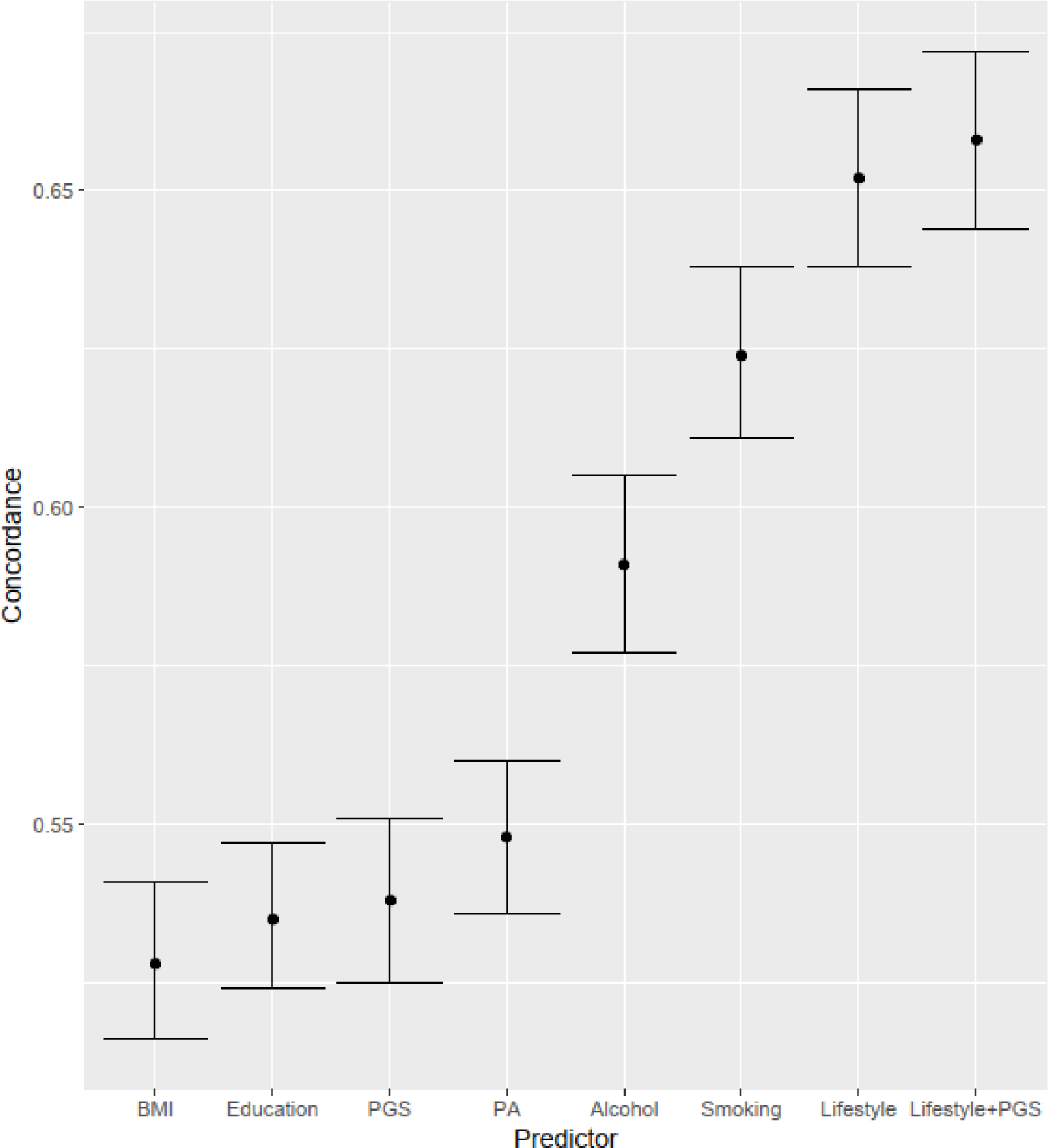
Concordance indices for PGS HGS and lifestyle factors. C-indices are from univariate Cox regression models for all-cause mortality in men with complete case data. A model with PGS HGS additionally adjusted for 10 genetic principal components of ancestry. PGS HGS, the polygenic score for hand grip strength; BMI, body mass index; PA, physical activity.

**Figure 4.**
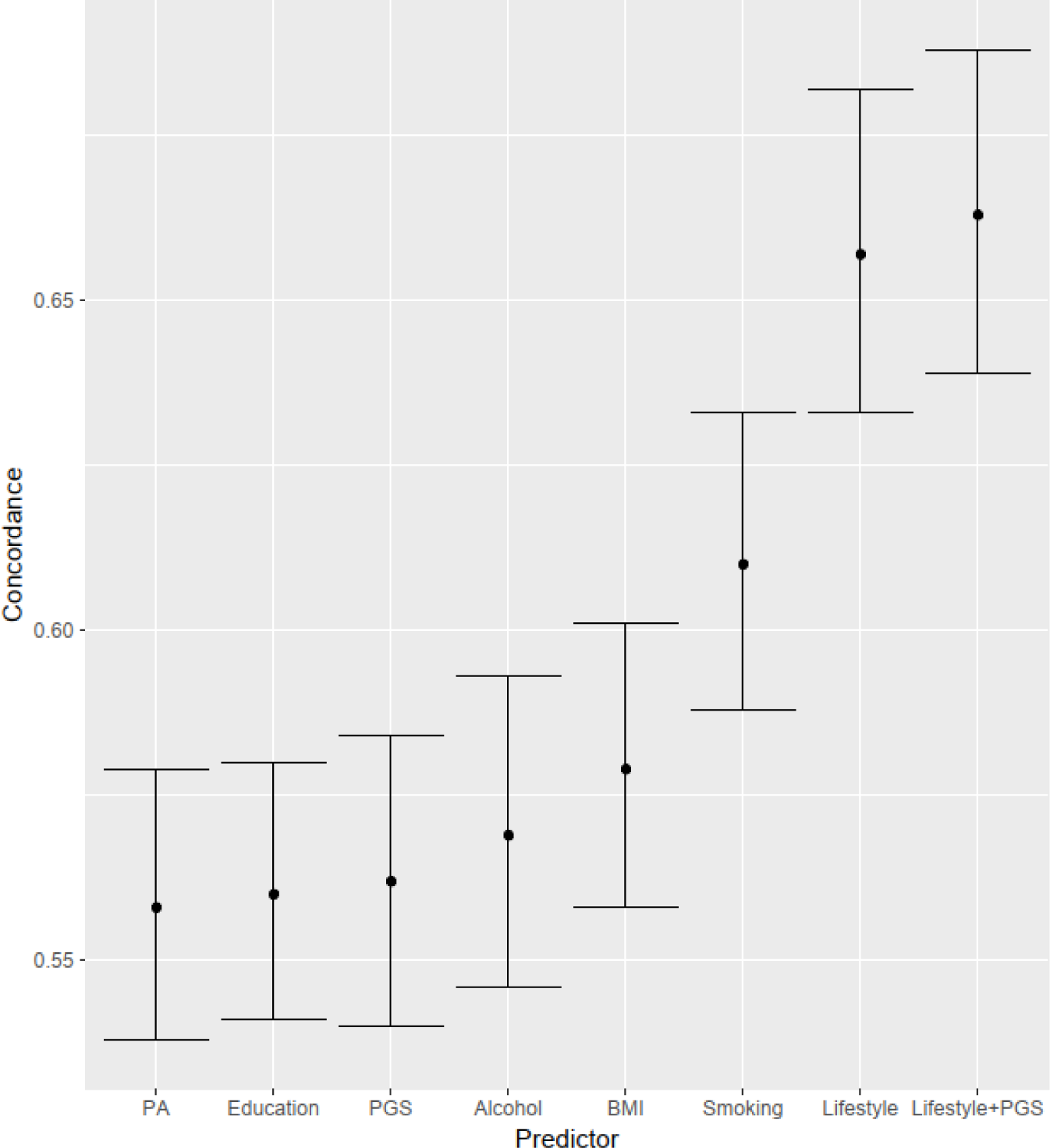
Concordance indices for PGS HGS and lifestyle factors. C-indices are from univariate Cox regression models for CVD mortality in men with complete case data. A model with PGS HGS additionally adjusted for 10 genetic principal components of ancestry. PGS HGS, the polygenic score for hand grip strength; CVD, cardiovascular disease; PA, physical activity; BMI, body mass index.

### Sensitivity analysis

As seen in Tables S1–S5, deviations from the PH assumptions were small and the results persisted even when time dependence was considered. Furthermore, in subgroup analyses conducted for apparently healthy participants, the risk estimates remained relatively consistent for both sexes (Tables S7–S12). However, potential bias in the results should be acknowledged due to the constrained sample size.

## Discussion

In this study, we investigated whether PGS HGS can serve as a proxy for an individual’s intrinsic capacity to survive premature mortality and whether this association is influenced by long-term leisure-time PA over adulthood. We observed that the genotype supporting higher muscle strength was independently associated with a lower mortality risk, especially CVD mortality, in men. Long-term leisure-time PA did not potentiate this association, but it may offer independent benefits. However, the associations between long-term leisure-time PA and CVD mortality were relatively small and attenuated further when other lifestyle and socioeconomic covariates were considered.

While low muscle strength is an established risk factor for mortality, the mechanisms underlying this association have not been fully explored. The beneficial impacts of muscle strength on health might be rooted in its importance in controlling the body’s energy homeostasis and its potential to counteract chronic inflammation.^22,44^ Skeletal muscle is closely intertwined with cardiometabolic health, serving as the primary organ involved in regulating glucose metabolism and lipid profiles; this, in turn, improves insulin sensitivity, promotes favorable body composition, and maintains blood pressure levels.^22^ Moreover, strong muscles act as reservoirs of energy and endurance, enhancing resilience in the face of acute illnesses or medical procedures.^44^ The complex biological mechanisms governing the association between muscle strength and mortality are intricately regulated by an interplay of genes, epigenetic mechanisms, and environmental factors.^45^ Our results imply that the genetic predisposition for higher muscle strength may play a role in underlying this association. Our findings align with a previous GWAS that indicated a causal effect of HGS on coronary heart disease and atrial fibrillation,^15^ both leading causes of CVD mortality. In addition, several studies have indicated causal relationships between HGS and various markers of adiposity.^15,16,46^ Similar to muscle weakness, excess adiposity is associated with an increased risk of metabolic and cardiovascular complications.^47^ Based on these findings, we can confirm that skeletal muscle strength is fundamental in preventing cardiovascular events and helps sustain a longer and healthy life.

Our data suggest that a higher genetic predisposition for muscle strength predicts a longer lifespan only in men. It is known that individual characteristics, such as sex or genetic variation, influence both muscle strength and its relationship with PA.^48,49^ For instance, loss of muscle strength significantly limits mobility, especially in older women, who have a smaller strength reserve than men.^50,51^ On the other hand, while men typically exhibit greater strength during early adulthood, they experience a sharper decline in HGS after midlife compared to women until this sex-related discrepancy in slope diminishes with age.^52,53^ Presumably, this reflects a higher likelihood of women for developing chronic conditions but leading a longer life; in contrast, men tend to experience an increased incidence of acute conditions, such as ischemic heart disease and stroke, leading to shorter lifespan.^45,54^ It is believed that different environmental factors are involved in shaping the sex-specific patterns of muscle strength decline,^49^ but twin studies have also shown that heritability estimates for HGS throughout life are consistently higher in men (72%) than in women (47%).^54^ Although the gap between women and men in terms of muscle strength, risk behavior, disease prevalence, and mortality is widely acknowledged, the reported sex-related differences regarding the prognostic value of HGS^5,6^ and PGS HGS for mortality^20^, as well as in the genetic architecture of human traits, are relatively inconsequential.^55,56^ Therefore, further research is necessary to validate the sex-related differences found in this present study.

As opposed to our hypothesis, long-term leisure-time PA did not moderate the association between PGS HGS and mortality; i.e., we could not find any evidence supporting an interaction effect between the two. Generally, it is difficult to accurately detect G × E interactions as they require considerably large sample sizes.^18^ A previous study suggested that environmental factors contributing to HGS trajectories could be modified by genes such as apolipoprotein E (ApoE).^48^ ApoE has several haplotypes with varying frequencies across populations, which may manifest differently depending on the context and the extent of influence of environmental factors. Additionally, the impact of these haplotypes may fluctuate depending on individual-related factors, such as sex, ethnicity, and age,^57^ and individual differences in environmental characteristics might be partly explained by hereditary factors.^58,59^ Therefore, interpreting G × E interactions might introduce bias if the environmental moderator is influenced by genetics itself, especially when the relevant genes can also directly impact the trait under scrutiny.^58^ Both muscle strength and PA are complex multifactorial traits, and it is believed that the same genetic background affecting PA partly overlaps with the mechanisms related to an individual’s metabolism.^60^ In our study, PGS HGS was not associated with PA, thus excluding the existence of a mediating pathway; however, genetic pleiotropy may exist between muscle strength and PA, indicating that both traits may be influenced, at least in part, by the same set of genes. Moreover, the PGSs used in our study were constructed considering only the main effects of genetic variants on HGS, ignoring possible interactions between each variant and environmental factors.^61^ Nonetheless, our results emphasize the importance of continued efforts to refine G × E modeling and suggest that, with further refinement, PGS HGS could become a valuable tool for advancing research in predicting health status in older adults and for implementing preventive strategies in individuals at risk.

Regular PA following recommended guidelines has been reported to improve health-related outcomes and decrease mortality risk in observational studies.^21,62^ However, in the present study, the independent associations between long-term leisure-time PA and mortality were relatively low. One explanation for this discrepancy might be that the changes in PA behavior over generations in previous population studies have focused mainly on the aerobic type and self-reported PA, although different forms of activity produce different adaptations.^63^ While aerobic exercise is linked to factors that can improve cardiometabolic health, such as cardiorespiratory fitness, blood lipid profiles, and hemodynamics, sustained commitment to muscle-strengthening exercises is particularly more relevant for preserving sufficient muscle strength and mass, components that are indeed essential in regulating glucose metabolism and basal metabolic rate.^27^ There is indeed evidence that long-term adherence to muscle-strengthening activities is associated with a lower risk of all-cause mortality, CVD, diabetes, and lung cancer independent of participation in aerobic activities.^28^ Although we observed no indication that sustained engagement in PA could moderate the association between PGS HGS and mortality, the genotype that supports higher muscle strength may interact with training response and training adherence.^64^ However, we were unable to investigate this matter in our study, so further research on this topic is warranted.

The present study has several strengths. First, although using twin cohort data may lead to volunteer bias or recruitment bias,^65^ data from the population-based older FTC cohort has shown to be representative of the general Finnish population, for example, in terms of overall mortality.^29^ Our study also included a long follow-up time of over 16 years with longitudinal lifestyle information and high-quality mortality data.^36,37^ Additionally, the PGS HGS utilized in this study is a reliable variable reflecting genetic predisposition to overall muscle strength and is based on a large number of genetic variants characteristic of European populations. Our study also had some limitations. First, PA was self-reported and based on surveys conducted in 1975, 1980, and 1990. Surveys can be dominated by socio-demographic trends typical of the era, with many popular aerobic activities perceived as masculine being over- represented in the questions, while the forms of PA used by women were not accurately surveyed.^29^ Second, the sample size in the older FTC cohort was relatively small for this type of analysis, which affected the power of our study results.^17^ Additionally, it limited our ability to analyze other causes of death. Third, UKBB participants are volunteers and may potentially exhibit better health and muscle strength than the broader population.^66^ Since our analysis was limited to individuals of European ancestry, this may restrict the generalizability of our findings. Due to the absence of a comparable dataset, we were unable to replicate our findings in another independent cohort. Finally, the current study design did not allow for the determination of a causal relationship between PGS HGS and mortality.

## Conclusions

Our findings indicate that genetic liability to higher muscle strength could potentially offer a slight shield against CVD mortality among males, irrespective of their PA levels later in life. The PGSs for muscle strength must be developed further and tested in a more diverse population; nevertheless, they can be a valuable tool for identifying individuals situated at the far ends of the genetic distribution of muscle strength and who are the most prone to cardiovascular events. Further research utilizing a larger sample size and varied study designs may help to unveil diverse avenues for integrating PGSs for muscle strength into clinical practice. Moreover, it could shed light on whether an individual’s inherent genetic propensity for muscle strength influences their response to exercise and capacity for improvement through training.

## Data Availability

All participants were informed about the study and their written consent was obtained. The full cohort data cannot be publicly available owing to the potential identifiability of the twin siblings in Finland. DNA samples and follow-up questionnaire data are available for the twins who had consented to transfer their data to the Biobank of the National Institute for Health and Welfare. These data are publicly available by following a standardized application procedure (https://thl.fi/en/research-and-development/thl-biobank/for-researchers/application-process). Full cohort data can be obtained from the Institute for Molecular Medicine Finland Data Access Committee (FIMM-DAC) by authorized researchers who have the appropriate IRB/ethics approval and an institutionally approved study plan. The GWAS summary statistics for maximum HGS were based on genetic and phenotypic data from the UK Biobank (UKBB) and derived from the publicly accessible website of the Pan-UKBB (https://pan.ukbb.broadinstitute.org/).

## Nonstandard Abbreviations and Acronyms

BMI: body mass index
CVD: cardiovascular disease
FTC: Finnish Twin Cohort
GWAS: genome-wide association study
HGS: hand grip strength
MET: metabolic equivalent
PA: physical activity
PC: principal component
PGS: polygenic score
UKBB: UK Biobank

## Acknowledgments

We extend our gratitude to the participants of the older FTC and UKBB studies for their valuable contributions to science and to the researchers who conducted the original data collection.

## References

1. López-Ortiz S, Lista S, Peñín-Grandes S, Pinto-Fraga J, Valenzuela PL, Nisticò R, Emanuele E, Lucia A, Santos-Lozano A. Defining and assessing intrinsic capacity in older people: a systematic review and a proposed scoring system. Ageing Res. Rev. 2022;79:101640.

2. Hughes VA, Frontera WR, Wood M, Evans WJ, Dallal GE, Roubenoff R, Fiatarone Singh MA. Longitudinal muscle strength changes in older adults: influence of muscle mass, physical activity, and health. J. Gerontol. A. Biol. Sci. Med. Sci. 2001;56:B209–217.

3. Cruz-Jentoft AJ, Baeyens JP, Bauer JM, Boirie Y, Cederholm T, Landi F, Martin FC, Michel J-P, Rolland Y, Schneider SM, et al. Sarcopenia: European consensus on definition and diagnosis: Report of the European Working Group on Sarcopenia in Older People. Age Ageing. 2010;39:412–423.

4. Damluji AA, Alfaraidhy M, AlHajri N, Rohant NN, Kumar M, Al Malouf C, Bahrainy S, Ji Kwak M, Batchelor WB, Forman DE, et al. Sarcopenia and cardiovascular diseases. Circulation. 2023;147:1534–1553.

5. García-Hermoso A, Cavero-Redondo I, Ramírez-Vélez R, Ruiz JR, Ortega FB, Lee D-C, Martínez-Vizcaíno V. Muscular strength as a predictor of all-cause mortality in an apparently healthy population: a systematic review and meta-analysis of data from approximately 2 million men and women. Arch. Phys. Med. Rehabil. 2018;99:2100–2113.e5.

6. Celis-Morales CA, Welsh P, Lyall DM, Steell L, Petermann F, Anderson J, Iliodromiti S, Sillars A, Graham N, Mackay DF, et al. Associations of grip strength with cardiovascular, respiratory, and cancer outcomes and all cause mortality: prospective cohort study of half a million UK Biobank participants. BMJ. 2018;361:k1651.

7. Koivunen K, Sillanpaa E, von Bonsdorff M, Sakari R, Tormakangas T, Rantanen T. Mortality risk among older people who did vs. did not sustain a fracture: baseline pre-fracture strength and gait speed as predictors in a 15-year follow-up. J. Gerontol. Biol. Sci. Med. Sci. 2019;

8. Heitmann BL, Frederiksen P. Thigh circumference and risk of heart disease and premature death: prospective cohort study. BMJ. 2009;339:b3292.

9. Rantanen T, Masaki K, Foley D, Izmirlian G, White L, Guralnik JM. Grip strength changes over 27 yr in Japanese-American men. J. Appl. Physiol. Bethesda Md 1985. 1998;85:2047–2053.

10. Henriksson H, Henriksson P, Tynelius P, Ortega FB. Muscular weakness in adolescence is associated with disability 30 years later: a population-based cohort study of 1.2 million men. Br. J. Sports Med. 2019;53:1221–1230.

11. Silventoinen K, Magnusson PKE, Tynelius P, Batty GD, Rasmussen F. Association of body size and muscle strength with incidence of coronary heart disease and cerebrovascular diseases: a population-based cohort study of one million Swedish men. Int. J. Epidemiol. 2009;38:110–118.

12. Timpka S, Petersson IF, Zhou C, Englund M. Muscle strength in adolescent men and risk of cardiovascular disease events and mortality in middle age: a prospective cohort study. BMC Med. 2014;12:62.

13. Kuh D, the New Dynamics of Ageing (NDA) Preparatory Network*. A life course approach to healthy aging, frailty, and capability. J. Gerontol. Ser. A. 2007;62:717–721.

14. Zempo H, Miyamoto-Mikami E, Kikuchi N, Fuku N, Miyachi M, Murakami H. Heritability estimates of muscle strength-related phenotypes: a systematic review and meta-analysis. Scand. J. Med. Sci. Sports. 2017;27:1537–1546.

15. Tikkanen E, Gustafsson S, Amar D, Shcherbina A, Waggott D, Ashley EA, Ingelsson E. Biological insights into muscular strength: genetic findings in the UK Biobank. Sci. Rep. 2018;8:6451.

16. Willems SM, Wright DJ, Day FR, Trajanoska K, Joshi PK, Morris JA, Matteini AM, Garton FC, Grarup N, Oskolkov N, et al. Large-scale GWAS identifies multiple loci for hand grip strength providing biological insights into muscular fitness. Nat. Commun. 2017;8:16015.

17. Choi SW, Mak TSH, O’Reilly PF. A guide to performing polygenic risk score analyses. Nat. Protoc. 2020;15:2759–2772.

18. Virolainen SJ, VonHandorf A, Viel KCMF, Weirauch MT, Kottyan LC. Gene– environment interactions and their impact on human health. Genes Immun. 2023;24:1– 11.

19. Herranen P, Palviainen T, Rantanen T, Tiainen K, Viljanen A, Kaprio J, Sillanpää E. A polygenic risk score for hand grip strength predicts muscle strength and proximal and distal functional outcomes among older women. Med. Sci. Sports Exerc. 2022;54:1889– 1896.

20. Herranen P, Koivunen K, Palviainen T, Kujala UM, Ripatti S, Kaprio J, Sillanpää E. Genome-wide polygenic score for muscle strength predicts risk for common diseases and lifespan: a prospective cohort study. J. Gerontol. Ser. A. 2024;79:glae064.

21. Singh R, Pattisapu A, Emery MS. US Physical activity guidelines: current state, impact and future directions. Trends Cardiovasc. Med. 2020;30:407–412.

22. Distefano G, Goodpaster BH. Effects of exercise and aging on skeletal muscle. Cold Spring Harb. Perspect. Med. 2018;8:a029785.

23. Rantanen T, Era P, Heikkinen E. Physical activity and the changes in maximal isometric strength in men and women from the age of 75 to 80 years. J. Am. Geriatr. Soc. 1997;45:1439–1445.

24. Sillanpaa E, Hakkinen A, Nyman K, Mattila M, Cheng S, Karavirta L, Laaksonen DE, Huuhka N, Kraemer WJ, Hakkinen K. Body composition and fitness during strength and/or endurance training in older men. Med. Sci. Sports Exerc. 2008;40:950–958.

25. Sillanpaa E, Hakkinen A, Punnonen K, Hakkinen K, Laaksonen DE. Effects of strength and endurance training on metabolic risk factors in healthy 40-65-year-old men. Scand. J. Med. Sci. Sports. 2009;19:885–895.

26. Cooper A, Lamb M, Sharp SJ, Simmons RK, Griffin SJ. Bidirectional association between physical activity and muscular strength in older adults: results from the UK Biobank study. Int. J. Epidemiol. 2017;46:141–148.

27. Brellenthin AG, Bennie JA, Lee D. Aerobic or muscle-strengthening physical activity: which is better for health? Curr. Sports Med. Rep. 2022;21:272–279.

28. Momma H, Kawakami R, Honda T, Sawada SS. Muscle-strengthening activities are associated with lower risk and mortality in major non-communicable diseases: a systematic review and meta-analysis of cohort studies. Br. J. Sports Med. 2022;56:755– 763.

29. Kaprio J, Bollepalli S, Buchwald J, Iso-Markku P, Korhonen T, Kovanen V, Kujala U, Laakkonen EK, Latvala A, Leskinen T, et al. The Older Finnish Twin Cohort - 45 years of follow-up. Twin Res. Hum. Genet. Off. J. Int. Soc. Twin Stud. 2019;22:240–254.

30. Hällfors J, Palviainen T, Surakka I, Gupta R, Buchwald J, Raevuori A, Ripatti S, Korhonen T, Jousilahti P, Madden PAF, et al. Genome-wide association study in Finnish twins highlights the connection between nicotine addiction and neurotrophin signaling pathway. Addict. Biol. 2019;24:549–561.

31. He L, Pitkäniemi J, Heikkilä K, Chou Y, Madden PAF, Korhonen T, Sarin A, Ripatti S, Kaprio J, Loukola A. Genome-wide time-to-event analysis on smoking progression stages in a family-based study. Brain Behav. 2016;6:e00462.

32. Howie BN, Donnelly P, Marchini J. A flexible and accurate genotype imputation method for the next generation of genome-wide association studies. PLoS Genet. 2009;5:e1000529.

33. Delaneau O, Howie B, Cox AJ, Zagury J-F, Marchini J. Haplotype estimation using sequencing reads. Am. J. Hum. Genet. 2013;93:687–696.

34. Lloyd-Jones LR, Zeng J, Sidorenko J, Yengo L, Moser G, Kemper KE, Wang H, Zheng Z, Magi R, Esko T, et al. Improved polygenic prediction by Bayesian multiple regression on summary statistics. Nat. Commun. 2019;10:5086.

35. International HapMap 3 Consortium, Altshuler DM, Gibbs RA, Peltonen L, Altshuler DM, Gibbs RA, Peltonen L, Dermitzakis E, Schaffner SF, Yu F, et al. Integrating common and rare genetic variation in diverse human populations. Nature. 2010;467:52–58.

36. Lahti RA, Penttilä A. The validity of death certificates: routine validation of death certification and its effects on mortality statistics. Forensic Sci. Int. 2001;115:15–32.

37. Lahti RA, Penttilä A. Cause-of-death query in validation of death certification by expert panel; effects on mortality statistics in Finland, 1995. Forensic Sci. Int. 2003;131:113–124.

38. Silventoinen K, Sarlio-Lähteenkorva S, Koskenvuo M, Lahelma E, Kaprio J. Effect of environmental and genetic factors on education-associated disparities in weight and weight gain: a study of Finnish adult twins. Am. J. Clin. Nutr. 2004;80:815–822.

39. Lehtovirta M, Pietiläinen KH, Levälahti E, Heikkilä K, Groop L, Silventoinen K, Koskenvuo M, Kaprio J. Evidence that BMI and type 2 diabetes share only a minor fraction of genetic variance: a follow-up study of 23,585 monozygotic and dizygotic twins from the Finnish Twin Cohort Study. Diabetologia. 2010;53:1314–1321.

40. Rintakoski K, Ahlberg J, Hublin C, Broms U, Madden P a. F, Könönen M, Koskenvuo M, Lobbezoo F, Kaprio J. Bruxism is associated with nicotine dependence: a nationwide Finnish twin cohort study. Nicotine Tob. Res. Off. J. Soc. Res. Nicotine Tob. 2010;12:1254–1260.

41. Carlsson S, Hammar N, Grill V, Kaprio J. Alcohol consumption and the incidence of type 2 diabetes: a 20-year follow-up of the Finnish twin cohort study. Diabetes Care. 2003;26:2785–2790.

42. Zhang Z, Reinikainen J, Adeleke KA, Pieterse ME, Groothuis-Oudshoorn CGM. Time-varying covariates and coefficients in Cox regression models. Ann. Transl. Med. 2018;6:121.

43. Kujala UM, Kaprio J, Koskenvuo M. Modifiable risk factors as predictors of all-cause mortality: the roles of genetics and childhood environment. Am. J. Epidemiol. 2002;156:985–993.

44. Argilés JM, Campos N, Lopez-Pedrosa JM, Rueda R, Rodriguez-Mañas L. Skeletal muscle regulates metabolism via interorgan crosstalk: roles in health and disease. J. Am. Med. Dir. Assoc. 2016;17:789–796.

45. Hägg S, Jylhävä J. Sex differences in biological aging with a focus on human studies. eLife. 10:e63425.

46. Ran S, Zhao M-F, Liu B-L. The causal association of sarcopenia with osteoporosis and obesity: a Mendelian randomization analysis. Osteoporos. Int. 2023;34:613–614.

47. Engin A. The definition and prevalence of obesity and metabolic syndrome. Adv. Exp. Med. Biol. 2017;960:1–17.

48. Petersen I, Pedersen NL, Rantanen T, Kremen WS, Johnson W, Panizzon MS, Christiansen L, Franz CE, McGue M, Christensen K, et al. G×E interaction influences trajectories of hand grip strength. Behav. Genet. 2016;46:20–30.

49. Sternäng O, Reynolds CA, Finkel D, Ernsth-Bravell M, Pedersen NL, Dahl Aslan AK. Factors associated with grip strength decline in older adults. Age Ageing. 2015;44:269– 274.

50. Rantanen T, Guralnik JM, Izmirlian G, Williamson JD, Simonsick EM, Ferrucci L, Fried LP. Association of muscle strength with maximum walking speed in disabled older women. Am. J. Phys. Med. Rehabil. 1998;77:299–305.

51. Rantanen T, Era P, Heikkinen E. Maximal isometric strength and mobility among 75- year-old men and women. Age Ageing. 1994;23:132–137.

52. Frederiksen H, Hjelmborg J, Mortensen J, Mcgue M, Vaupel J, Christensen K. Age trajectories of grip strength: cross-sectional and longitudinal data among 8,342 Danes aged 46 to 102. Ann. Epidemiol. 2006;16:554–562.

53. Nahhas RW, Choh AC, Lee M, Chumlea WMC, Duren DL, Siervogel RM, Sherwood RJ, Towne B, Czerwinski SA. Bayesian longitudinal plateau model of adult grip strength. Am. J. Hum. Biol. 2010;22:648–656.

54. Finkel D, Pedersen NL, Reynolds CA, Berg S, de Faire U, Svartengren M. Genetic and environmental influences on decline in biobehavioral markers of aging. Behav. Genet. 2003;33:107–123.

55. Flynn E, Tanigawa Y, Rodriguez F, Altman RB, Sinnott-Armstrong N, Rivas MA. Sex-specific genetic effects across biomarkers. Eur. J. Hum. Genet. 2021;29:154–163.

56. Bernabeu E, Canela-Xandri O, Rawlik K, Talenti A, Prendergast J, Tenesa A. Sex differences in genetic architecture in the UK Biobank. Nat. Genet. 2021;53:1283–1289.

57. Abondio P, Bruno F, Luiselli D. Apolipoprotein E (APOE) Haplotypes in healthy subjects from worldwide macroareas: a population genetics perspective for cardiovascular disease, neurodegeneration, and dementia. Curr. Issues Mol. Biol. 2023;45:2817–2831.

58. Vinkhuyzen A a. E, Van Der Sluis S, De Geus EJC, Boomsma DI, Posthuma D. Genetic influences on ‘environmental’ factors. Genes Brain Behav. 2010;9:276–287.

59. de Geus EJC. Genetic pathways underlying individual differences in regular physical activity. Exerc. Sport Sci. Rev. 2023;51:2–18.

60. Sillanpää E, Palviainen T, Ripatti S, Kujala UM, Kaprio J. Polygenic score for physical activity is associated with multiple common diseases. Med. Sci. Sports Exerc. 2022;54:280–287.

61. Tang Y, You D, Yi H, Yang S, Zhao Y. IPRS: Leveraging gene-environment interaction to reconstruct polygenic risk score. Front. Genet. 2022;13:801397.

62. Zhao G, Li C, Ford ES, Fulton JE, Carlson SA, Okoro CA, Wen XJ, Balluz LS. Leisure-time aerobic physical activity, muscle-strengthening activity and mortality risks among US adults: the NHANES linked mortality study. Br. J. Sports Med. 2014;48:244–249.

63. Sylvia LG, Bernstein EE, Hubbard JL, Keating L, Anderson EJ. A practical guide to measuring physical activity. J. Acad. Nutr. Diet. 2014;114:199–208.

64. Jones N, Kiely J, Suraci B, Collins D, de Lorenzo D, Pickering C, Grimaldi K. A genetic-based algorithm for personalized resistance training. Biol. Sport. 2016;33:117–126.

65. Tambs K, Rønning T, Prescott CA, Kendler KS, Reichborn-Kjennerud T, Torgersen S, Harris JR. The Norwegian Institute of Public Health twin study of mental health: examining recruitment and attrition bias. Twin Res. Hum. Genet. Off. J. Int. Soc. Twin Stud. 2009;12:158–168.

66. Fry A, Littlejohns TJ, Sudlow C, Doherty N, Adamska L, Sprosen T, Collins R, Allen NE. Comparison of sociodemographic and health-related characteristics of uk biobank participants with those of the general population. Am. J. Epidemiol. 2017;186:1026–1034.

