## Supplemental material for "Genetic Liability to Higher Muscle Strength Associates with a Lower Risk of Cardiovascular Disease Mortality in Men Irrespective of Physical Activity in Adulthood: A Longitudinal Cohort Study"

As seen in Table S1, in men, alcohol consumption violated the PH assumption in the model predicting all-cause mortality (P=0.018 for the scaled Schoenfeld residuals test), and BMI violated the PH assumption in the model predicting CVD mortality (P=0.047). In women, the PH assumption was violated in the model where PGS HGS was used to predict CVD mortality (P=0.018; Table S2). After treating these variables as time-dependent covariates in the stratified analyses by age intervals (Tables S3–S5), PH assumptions were corrected (*P*=0.494, *P*=0.440, and *P*=0.420, respectively).

Table S1. Test for Violation of the Assumption of Proportional Hazards: All-Cause and CVD Mortality in Men

| | All-cause mortality | CVD mortality |
|---|---|---|
| Variable | *P* value | *P* value |
| PGS HGS | 0.109 | 0.527 |
| Physical activity | 0.766 | 0.765 |
| Education | 0.770 | 0.801 |
| BMI | 0.134 | 0.047 |
| Smoking | 0.330 | 0.601 |
| Alcohol consumption | 0.018 | 0.112 |
| GLOBAL | 0.096 | 0.322 |

Results for the scaled Schoenfeld residuals test. CVD, cardiovascular related; PGS HGS, polygenic score for hand grip strength; BMI, body mass index.

Table S2. Test for Violation of the Assumption of Proportional Hazards: All-Cause and CVD Mortality in Women

| | All-cause mortality | CVD mortality |
|---|---|---|
| Variable | *P* value | *P* value |
| PGS HGS | 0.540 | 0.018 |
| Physical activity | 0.700 | 0.552 |
| Education | 0.960 | 0.771 |
| BMI | 0.560 | 0.268 |
| Smoking | 0.330 | 0.367 |
| Alcohol consumption | 0.890 | 0.484 |
| GLOBAL | 0.750 | 0.226 |

Results for the scaled Schoenfeld residuals test. CVD, cardiovascular related; PGS HGS, polygenic score for hand grip strength; BMI, body mass index.

Table S3. Association of PGS HGS with All-Cause Mortality in Men

| **All-cause mortality** | **Model 1** | ***P* value** | **Model 2** | ***P* value** | **Model 3** | ***P* value** |
|---|---|---|---|---|---|---|
| Cases / N | 1550 / 4148 | | 807 / 2522 | | 772 / 2399 | |
| PGS HGS (ZScore) | 0.93 (0.89 to 0.98) | 0.004 | 0.92 (0.86 to 0.98) | 0.009 | 0.94 (0.88 to 1.01) | 0.090 |
| Physical activity (MET h/day) | | | 0.93 (0.90 to 0.96) | <0.001 | 0.96 (0.92 to 0.99) | 0.009 |
| Education (years) | | | | | 0.97 (0.94 to 0.99) | 0.009 |
| BMI (kg/m$^2$) | | | | | 1.03 (1.01 to 1.06) | 0.017 |
| Smoking | | | | | | |
| Never | | | | | ref | |
| Occasional | | | | | 1.22 (0.81 to 1.85) | 0.310 |
| Former | | | | | 1.34 (1.11 to 1.61) | 0.002 |
| Light | | | | | 1.89 (1.28 to 2.80) | 0.001 |
| Medium | | | | | 2.20 (1.73 to 2.78) | <0.001 |
| Heavy | | | | | 3.30 (2.61 to 4.17) | <0.001 |
| Alcohol consumption (g/day) | | | | | | |
| ≤ 65 years | | | | | 1.01 (1.01 to 1.02) | <0.001 |
| 65–75 years | | | | | 1.01 (1.00 to 1.01) | 0.002 |
| 75–85 years | | | | | 1.01 (1.00 to 1.02) | 0.010 |
| >85 years | | | | | 1.00 (0.98 to 1.02) | 0.957 |

Multivariable Cox regression analysis; Values are Hazard Ratios with 95% Confidence intervals; All models are additionally adjusted for 10 principal components and family relatedness; PGS HGS, the polygenic score for handgrip strength; MET, metabolic equivalent of task; BMI, body mass index.

Table S4. Associations of PGS HGS with CVD Mortality in Men

| **CVD mortality** | **Model 1** | ***P* value** | **Model 2** | ***P* value** | **Model 3** | ***P* value** |
|---|---|---|---|---|---|---|
| **Cases / N** | 589 / 4148 | | 289 / 2522 | | 277 / 2399 | |
| PGS HGS (ZScore) | 0.88 (0.81 to 0.96) | 0.003 | 0.85 (0.76 to 0.96) | 0.008 | 0.85 (0.75 to 0.96) | 0.008 |
| Physical activity (MET h/day) | | | 0.93 (0.88 to 0.99) | 0.016 | 0.96 (0.90 to 1.01) | 0.138 |
| Education (years) | | | | | 0.94 (0.89 to 0.98) | 0.004 |
| BMI (kg/m$^2$) | | | | | 1.07 (1.02 to 1.11) | 0.002 |
| ≤ 65 years | | | | | 1.11 (1.01 to 1.23) | 0.037 |
| 65–75 years | | | | | 1.10 (1.04 to 1.16) | 0.001 |
| 75–85 years | | | | | 1.02 (0.95 to 1.08) | 0.623 |
| >85 years | | | | | 1.07 (0.92 to 1.26) | 0.380 |
| Smoking | | | | | | |
| Never | | | | | ref | |
| Occasional | | | | | 1.15 (0.59 to 2.25) | 0.681 |
| Former | | | | | 1.58 (1.15 to 2.17) | 0.005 |
| Light | | | | | 1.68 (0.86 to 3.27) | 0.130 |
| Medium | | | | | 1.95 (1.25 to 3.05) | 0.003 |
| Heavy | | | | | 3.27 (2.16 to 4.97) | <0.001 |
| Alcohol consumption | | | | | 1.01 (1.00 to 1.01) | 0.016 |

Multivariable Cox regression analysis; Values are Hazard Ratios with 95% Confidence intervals; All models are additionally adjusted for 10 principal components and family relatedness; CVD, cardiovascular disease; PGS HGS, the polygenic score for handgrip strength; MET, metabolic equivalent of task; BMI, body mass index.

Table S5. Associations of PGS HGS with CVD Mortality in Women

| **CVD mortality** | **Model 1** | ***P* value** | **Model 2** | ***P* value** | **Model 3** | ***P* value** |
|---|---|---|---|---|---|---|
| Cases / N | 500 / 4667 | | 172 / 3054 | | 171 / 3012 | |
| PGS HGS (ZScore) | | | | | | |
| ≤ 65 years | 1.00 (0.62 to 1.61) | 0.991 | 1.11 (0.62 to 1.99) | 0.723 | 1.06 (0.59 to 1.93) | 0.837 |
| 65–75 years | 1.25 (1.00 to 1.55) | 0.051 | 1.27 (0.97 to 1.67) | 0.087 | 1.25 (0.94 to 1.66) | 0.126 |
| 75–85 years | 0.96 (0.83 to 1.12) | 0.626 | 1.08 (0.86 to 1.37) | 0.510 | 1.09 (0.86 to 1.39) | 0.474 |
| >85 years | 0.87 (0.76 to 0.99) | 0.037 | 0.74 (0.56 to 0.99) | 0.045 | 0.71 (0.52 to 0.99) | 0.041 |
| Physical activity (MET h/day) | | | 0.95 (0.86 to 1.05) | 0.329 | 1.00 (0.91 to 1.10) | 0.987 |
| Education (years) | | | | | 0.96 (0.90 to 1.02) | 0.210 |
| BMI (kg/m$^2$) | | | | | 1.08 (1.05 to 1.12) | <0.001 |
| Smoking | | | | | | |
| Never | | | | | ref | |
| Occasional | | | | | 1.64 (0.75 to 3.58) | 0.216 |
| Former | | | | | 0.82 (0.46 to 1.47) | 0.507 |
| Light | | | | | 1.08 (0.47 to 2.45) | 0.863 |
| Medium | | | | | 2.50 (1.44 to 4.36) | 0.001 |
| Heavy | | | | | 3.84 (1.79 to 8.27) | 0.001 |
| Alcohol consumption g/day | | | | | 0.97 (0.93 to 1.01) | 0.120 |

Multivariable Cox regression analysis; Values are Hazard Ratios with 95% Confidence intervals; All models are additionally adjusted for 10 principal components and family relatedness; CVD, cardiovascular disease; PGS HGS, the polygenic score for handgrip strength; MET, metabolic equivalent of task; BMI, body mass index.

Table S6. Interactions Between PGS HGS and Physical Activity

| | **Men** | | **Women** | |
|---|---|---|---|---|
| *All-cause mortality* | | *P* value | | *P* value |
| Cases / N | 772 / 2399 | | 593 / 3012 | |
| PGS HGS*Physical activity | 1.01 (0.98 to 1.04) | 0.370 | 1.04 (0.99 to 1.09) | 0.109 |
| *CVD mortality* | | | | |
| Cases / N | 277 / 2399 | | 171 / 3012 | |
| PGS HGS*Physical activity | 1.03 (0.98 to 1.09) | 0.257 | 1.11 (1.02 to 1.20) | 0.011 |

Multivariable Cox regression analysis; Values are Hazard Ratios with 95% Confidence intervals; All models are additionally adjusted for 10 principal components and family relatedness; PGS HGS, the polygenic score for handgrip strength; CVD, cardiovascular disease.

Table S7. Test for Violation of the Assumption of Proportional Hazards: All-Cause and CVD Mortality Among Apparently Healthy Men

| | All-cause mortality | CVD mortality |
|---|---|---|
| Variable | *P* value | *P* value |
| PGS HGS | 0.140 | 0.590 |
| Physical activity | 0.230 | 0.770 |
| Education | 0.340 | 0.970 |
| BMI | 0.210 | 0.110 |
| Smoking | 0.830 | 0.550 |
| Alcohol consumption | 0.170 | 0.310 |
| GLOBAL | 0.390 | 0.520 |

Results for the scaled Schoenfeld residuals test. CVD, cardiovascular related; PGS HGS, polygenic score for hand grip strength; BMI, body mass index.

Table S8. Test for Violation of the Assumption of Proportional Hazards: All-Cause and CVD Mortality Among Apparently Healthy Women

| | All-cause mortality | CVD mortality |
|---|---|---|
| Variable | *P* value | *P* value |
| PGS HGS | 0.480 | 0.091 |
| Physical activity | 0.740 | 0.894 |
| Education | 0.950 | 0.708 |
| BMI | 0.750 | 0.147 |
| Smoking | 0.570 | 0.301 |
| Alcohol consumption | 0.690 | 0.707 |
| GLOBAL | 0.900 | 0.310 |

Results for the scaled Schoenfeld residuals test. CVD, cardiovascular related; PGS HGS, polygenic score for hand grip strength; BMI, body mass index.

Table S9. Association of PGS HGS with All-Cause Mortality Among Apparently Healthy Men

| **All-cause mortality** | **Model 1** | ***P* value** | **Model 2** | ***P* value** | **Model 3** | ***P* value** |
|---|---|---|---|---|---|---|
| Cases / N | 980 / 3041 | | 638 / 2175 | | 605 / 2063 | |
| PGS HGS (ZScore) | 0.92 (0.87 to 0.98) | 0.007 | 0.91 (0.84 to 0.97) | 0.005 | 0.94 (0.87 to 1.01) | 0.075 |
| Physical activity (MET h/day) | | | 0.94 (0.91 to 0.98) | 0.001 | 0.97 (0.93 to 1.00) | 0.079 |
| Education (years) | | | | | 0.96 (0.94 to 0.99) | 0.014 |
| BMI (kg/m$^2$) | | | | | 1.02 (0.99 to 1.05) | 0.139 |
| Smoking | | | | | | |
| Never | | | | | ref | |
| Occasional | | | | | 1.22 (0.78 to 1.91) | 0.329 |
| Former | | | | | 1.18 (0.95 to 1.47) | 0.137 |
| Light | | | | | 1.96 (1.27 to 3.02) | 0.002 |
| Medium | | | | | 2.23 (1.71 to 2.91) | <0.001 |
| Heavy | | | | | 3.20 (2.45 to 4.17) | <0.001 |
| Alcohol consumption (g/day) | | | | | 1.01 (1.01to 1.02) | <0.001 |

Multivariable Cox regression analysis; Values are Hazard Ratios with 95% Confidence intervals; All models are additionally adjusted for 10 principal components and family relatedness; PGS HGS, polygenic score for handgrip strength; MET, metabolic equivalent of task; BMI, body mass index.

Table S10. Association of PGS HGS with CVD Mortality Among Apparently Healthy Men

| **CVD mortality** | **Model 1** | ***P* value** | **Model 2** | ***P* value** | **Model 3** | ***P* value** |
|---|---|---|---|---|---|---|
| Cases / N | 346 / 3041 | | 216 / 2175 | | 205 / 2063 | |
| PGS HGS (ZScore) | 0.91 (0.82 to 1.01) | 0.067 | 0.87 (0.76 to 1.00) | 0.043 | 0.87 (0.76 to 1.00) | 0.057 |
| Physical activity (MET h/day) | | | 0.96 ( 0.91 to 1.02) | 0.161 | 0.98 (0.92 to 1.05) | 0.586 |
| Education (years) | | | | | 0.92 (0.88 to 0.98) | 0.004 |
| BMI (kg/m$^2$) | | | | | 1.06 (1.01 to 1.11) | 0.018 |
| Smoking | | | | | | |
| Never | | | | | ref | |
| Occasional | | | | | 1.02 (0.52 to 2.02) | 0.947 |
| Former | | | | | 1.17 (0.81 to 1.70) | 0.407 |
| Light | | | | | 1.49 (0.68 to 3.24) | 0.315 |
| Medium | | | | | 1.98 (1.22 to 3.24) | 0.006 |
| Heavy | | | | | 2.77 (1.68 to 4.57) | <0.001 |
| Alcohol consumption (g/day) | | | | | 1.01 (1.01 to 1.02) | 0.001 |

Multivariable Cox regression analysis; Values are Hazard Ratios with 95% Confidence intervals; All models are additionally adjusted for 10 principal components and family relatedness; CVD, cardiovascular disease; PGS HGS, polygenic score for handgrip strength; MET, metabolic equivalent of task; BMI, body mass index.

Table S11. Association of PGS HGS with All-Cause Mortality Among Apparently Healthy Women

| **All-cause mortality** | **Model 1** | ***P* value** | **Model 2** | ***P* value** | **Model 3** | ***P* value** |
|---|---|---|---|---|---|---|
| Cases / N | 754 / 3318 | | 438 / 2503 | | 434 / 2469 | |
| PGS HGS (ZScore) | 1.05 (0.97 to 1.12) | 0.219 | 1.10 (1.00 to 1.21) | 0.043 | 1.10 (1.01 to 1.21) | 0.036 |
| Physical activity (MET h/day) | | | 1.02 (0.97 to 1.07) | 0.548 | 1.02 (0.97 to 1.07) | 0.414 |
| Education (years) | | | | | 0.98 (0.95 to 1.02) | 0.318 |
| BMI (kg/m$^2$) | | | | | 1.02 (1.00 to 1.05) | 0.063 |
| Smoking | | | | | | |
| Never | | | | | ref | |
| Occasional | | | | | 1.51 (0.83 to 2.77) | 0.178 |
| Former | | | | | 1.25 (0.93 to 1.69) | 0.132 |
| Light | | | | | 1.29 (0.82 to 2.05) | 0.273 |
| Medium | | | | | 2.18 (1.58 to 3.01) | <0.001 |
| Heavy | | | | | 3.63 (2.36 to 5.57) | <0.001 |
| Alcohol consumption (g/day) | | | | | 1.01 (1.01 to 1.02) | <0.001 |

Multivariable Cox regression analysis; Values are Hazard Ratios with 95% Confidence intervals; All models are additionally adjusted for 10 principal components and family relatedness; PGS HGS, polygenic score for handgrip strength; MET, metabolic equivalent of task; BMI, body mass index.

Table S12. Association of PGS HGS with CVD Mortality Among Apparently Healthy Women

| **CVD mortality** | **Model 1** | ***P* value** | **Model 2** | ***P* value** | **Model 3** | ***P* value** |
|---|---|---|---|---|---|---|
| Cases / N | 241 / 3318 | | 115 / 2503 | | 114 / 2469 | |
| PGS HGS (ZScore) | 0.97 (0.86 to 1.09) | 0.578 | 1.04 (0.88 to 1.23) | 0.657 | 1.04 (0.87 to 1.24) | 0.674 |
| Physical activity (MET h/day) | | | 0.99 (0.89 to 1.10) | 0.902 | 1.03 (0.93 to 1.14) | 0.598 |
| Education (years) | | | | | 0.97 (0.91 to 1.04) | 0.425 |
| BMI (kg/m$^2$) | | | | | 1.08 (1.04 to 1.12) | <0.001 |
| Smoking | | | | | | |
| Never | | | | | ref | |
| Occasional | | | | | 2.16 (0.74 to 6.27) | 0.157 |
| Former | | | | | 0.73 (0.34 to 1.56) | 0.411 |
| Light | | | | | 0.93 (0.35 to 2.45) | 0.876 |
| Medium | | | | | 2.69 (1.39 to 5.19) | 0.003 |
| Heavy | | | | | 4.45 (1.76 to 11.28) | 0.002 |
| Alcohol consumption (g/day) | | | | | 0.99 (0.95 to 1.02) | 0.497 |

Multivariable Cox regression analysis; Values are Hazard Ratios with 95% Confidence intervals; All models are additionally adjusted for 10 principal components and family relatedness; CVD, cardiovascular disease; PGS HGS, polygenic score for handgrip strength; MET, metabolic equivalent of task; BMI, body mass index.
